# Omega-3 Index is Directly Associated with a Healthy Red Blood Cell Distribution Width

**DOI:** 10.1101/2021.10.22.21264652

**Authors:** Michael I. McBurney, Nathan L. Tintle, William S. Harris

## Abstract

Low red blood cell (RBC) membrane content of EPA and DHA, i.e., the omega-3 index (O3I), and elevated RBC distribution width (RDW) are risk factors for all-cause mortality. O3I and RDW are related with membrane fluidity and deformability. Our objective was to determine if there is a relationship between O3I and RDW in healthy adults. Subjects without inflammation or anemia, and with values for O3I, RDW, high-sensitivity C-reactive protein (CRP), body mass index (BMI), age and sex were identified (n=25,485) from a clinical laboratory dataset of >45,000 individuals. RDW was inversely associated with O3I <u>in both sexes</u> before and after (both p<0.00001) adjusting models for sex, age, BMI and CRP. Stratification by sex revealed a sex-O3I interaction with the RDW-O3I slope (p<0.00066) being especially steep in females with O3I ≤5.6%. In healthy adults of both sexes, the data suggested that an O3I of >5.6% may help maintain normal RBC structural and functional integrity.

## 1. Introduction

Mean red blood cell (RBC) corpuscular volume (MCV) and reticulocyte count were the principal metrics traditionally used to identify anemic disorders until the advent of automated blood cell analyzers that could routinely measure the types and distribution of blood cells. Among these metrics is the RBC distribution width (RDW) [1] which reflects RBC size heterogeneity. High RDW values are associated with decreased deformability [2] and RDW is used to diagnose sickle cell [3] and nutritional anemias (iron, folic acid and vitamin B12) [1,4,5]. In recent years, RDW has been found to be a predictor of multiple adverse health outcomes unrelated to anemia, including increased risk for death from cardiovascular disease [6–12], SARS-CoV-2 [13–15], sepsis [16,17], lung disease [18], and cancer [19,20].

Like all cell membranes, RBC membranes are a complex lipid bilayer [21]. Fatty acids <u>esterified in phospholipids</u> contribute to the structural integrity and function of cell membranes. In particular, the membrane content of omega-3 fatty acids, EPA and DHA, the sum of which is called the omega-3 index (O3I) [22], increases with omega-3 fatty acid intake [23,24] and affects cell deformability [25–28]. The O3I is a stable biomarker of long term EPA and DHA intake [24,29]. Since both a low O3I [30] and an elevated RDW [7,31,32] are known to be associated with risk for earlier all-cause mortality (**Supplemental Table 1**), the aim of this cross-sectional analysis was to determine if there was an inverse relationship between the O3I and RDW in healthy adults without evidence of inflammation or tissue injury. If there was, then the former may play a role in maintaining a healthy distribution of RBC sizes in adults. <u>Furthermore, such a relationship could support the identification of a target or healthy O3I. By establishing a nutrient: structure-function relationship in healthy individuals, these data could be used as the basis for establishing a Dietary Reference Intake (DRI) for the long-chain omega-3 fatty acids, i.e.</u>, <u>EPA+DHA</u>.

## 2. Patients and Methods

This was a cross-sectional analysis of data from blood samples submitted for testing to Health Diagnostic Laboratory, Inc (HDL, Inc., Richmond VA) as part of routine clinical assessment between 2011-2012. Data on RBC structural characteristics, i.e., MCV and RDW, along with hemoglobin (Hb), high-sensitivity C-reactive protein (CRP)<u>, arachidonic acid (AA), EPA, DHA,</u> and O3I were extracted from 45,715 adults (≥18 years) without any linked patient identifiers except age, sex and BMI (**Supplemental Table 2**). Next, 598 individuals with <u>extreme</u> O3I values <u>(i.e., <2.15</u> and <u>>11.5%, the upper and lower</u> 0.5 <u>percentiles of the cohort</u>), were excluded (n=45,257). Since inflammation affects erythropoiesis, modulates RBC life span, and increases RDW [33], 14,329 individuals with chronic inflammation (CRP >3mg/L) [34–38] were excluded. Similarly, anemia also increases RDW [1,39], and so individuals with MCV >100 fL and low Hb (Hb <13 g/dL for men and Hb <12 g/dL for women [40] were also excluded (**Figure 1**).

**Figure 1.**
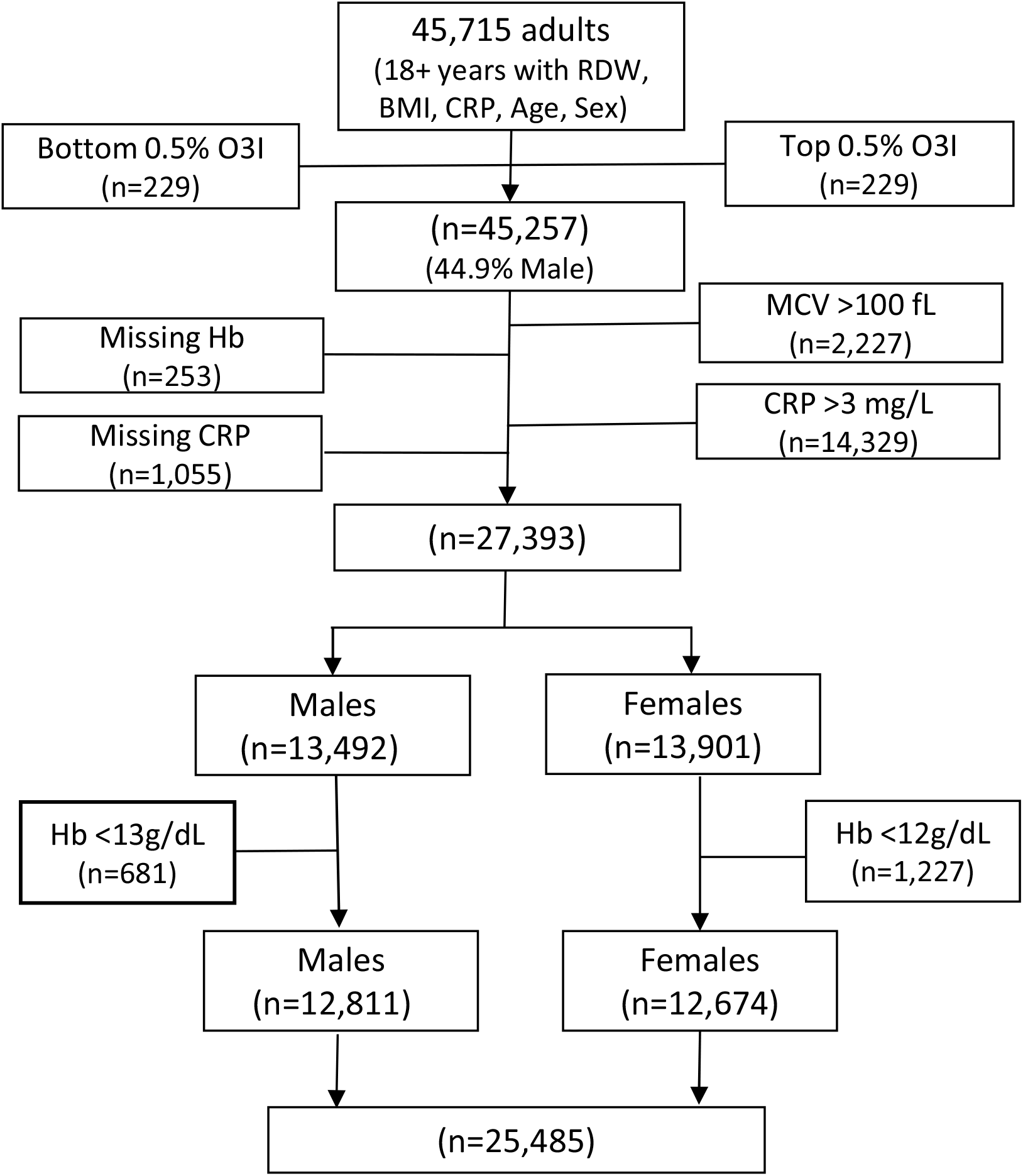
Analytical sample flow chart. RDW, red blood cell distribution width; O3I, omega-3 index; MCV, mean corpuscular cell volume; Hb, hemoglobin.

This resulted in a final, analytical dataset of 25,485 individuals (49.7% female) without laboratory evidence of anemia or active infection, tissue injury, or inflammation (**Table 1**).

**Table 1.** Characteristics of the study population. Mean±SD.

| Table 1. Characteristics of the study population. Mean±SD. |  |  |  |  |
| --- | --- | --- | --- | --- |
| Variable<sup>1</sup> | All<br>(n=25,485) | Males<br>(n=12,811) | Females<br>(n=12,674) | Male vs Female<br>Adjusted<sup>2</sup><br>p-value |
| O3I (%) | 4.99±1.83 | 4.90±1.81 | 5.07±1.85 | 0.0003 |
| RDW (%) | 13.60±0.90 | 13.60±0.83 | 13.69±0.96 | 0.0002 |
| MCV (fL) | 92.45±4.27 | 92.33±4.25 | 92.57±4.28 | 0.36 |
| Hb (g/dL) | 14.40±1.21 | 15.14±1.03 | 13.65±0.88 | <0.00001 |
| CRP (mg/L) | 1.23±0.77 | 1.21±0.75 | 1.25±0.78 | <0.00001 |
| BMI (kg/m<sup>2</sup>) | 27.7±5.3 | 28.8±4.8 | 25.6±5.5 | <0.00001 |
| Age (years) | 54.3±14.8 | 54.1±14.4 | 54.4±15.2 | 0.077 |
| <sup>1</sup>O3I, Omega-3 index; RDW, red blood cell distribution width; MCV, mean corpuscular volume; Hb, hemoglobin; CRP, high-sensitivity C-reactive protein; BMI, body mass index.<br><sup>2</sup>Models were adjusted for age, BMI, and CRP (except when the model was predicting these variables). |  |  |  |  |

### 2.1 Laboratory methods

Blood samples were drawn after an overnight fast and shipped with cold packs to HDL, Inc. for analysis with a Beckman-Coulter DxH 800 hematology analyzer (Brea, CA, USA). Samples were prepared at each clinical site according to standardized instructions as previously described [41]. RDW is the SD of the RBC volume divided by the MCV multiplied by 100 [39]. For fatty acid analysis, RBCs were separated from plasma by centrifugation and analyzed using gas chromatography as previously described [42]. The University of South Dakota Institutional Review Board reviewed and approved the analysis of deidentified HLD, Inc. laboratory data (IRB-21-147).

### 2.2 Statistical methods

Sample characteristics were summarized using standard statistical methods (e.g., means, SDs, correlations) with t-tests or adjusted linear models used to compare characteristics of male and female participants above and below O3I values that were ultimately identified as primary cut points in the RDW-O3I curves (see below). Splines were fit using 3^rd^ degree polynomials with knots at each decile in R (version 3.6.2; splines package). Unadjusted models used a linear model to predict RDW values by splines of O3I. Adjusted models accounted for sex, age, BMI and CRP values in the linear models accounting for potential non-linear relationships using splines.

In order to identify significant changes in the shape of the RDW-O3I relationship, we used a “sliding O3I window” approach. The width of each window was three O3I percentage points (e.g., 3% to 6%). By moving the window up by 0.1% increments and repeatedly testing for significant differences between the mean RDW in the lower vs. the upper half of the window, we sought to discover O3I cut points where the RDW-O3I relationship appeared to flatten. These would be O3I values above which the “effect” of an increase in O3I on RDW had little impact. We began with a window midpoint of O3I =2.6%, and we used adjusted linear models in R to test for upper vs lower half differences. We identified the first window which did not have a statistically significant difference in upper and lower mean RDW values. The midpoint of this window was chosen as the O3I cut point to be used in further analysis. To evaluate the potential moderating effect of sex on the O3I relationship with RDW, MCV, Hb, CRP, BMI and age, we inserted an interaction term into the relevant, adjusted model. Pearson correlations were used to assess strength and direction of linear association between covariates and RDW. Statistical significance was set to 0.05 for all analyses and 95% confidence bands are provided where appropriate.

## 3. Results

The final dataset included 25,485 individuals (**Figure 1**) without acute inflammation or anemia. Mean BMI, O3I, MCV, Hb, and CRP values differed significantly between males and females in adjusted models (**Table 1**). RDW was significantly (p<0.00001) and inversely associated with O3I in unadjusted (**Figure 2A**) and adjusted models (p<0.00001) (**Figure 2B**). <u>Independently, EPA and DHA had similar ‘hockey stick’ RDW-fatty acid relationships with the bases of the curves being ∼3% and ∼7.3%, respectively (</u>**<u>Supplemental Figure 2</u>**<u>). RDW was not significantly related with AA (</u>**<u>Supplemental Figure 2</u>**<u>, r=0.06).</u> Pearson correlations (r) with RDW were r=0.21 (age), r=0.11 (CRP), r=0.14 (BMI), r=-0.25 (MCV), and r=-0.12 (Hb). The O3I cut point determined in the sliding window analysis, i.e. when the lower and upper half of an “O3I sliding window” yielded mean RDW values that did not significantly differ, was 5.6% (**Figure 2B**). That is, below an O3I of 5.6%, the curve was clearly steeper than it was above 5.6%. Because of significant sex differences in RDW and O3I in the adjusted model (**Table 1**), and because of the clear differences in the shapes of the RDW-O3I relationship between men and women **(Figure 2)**, we stratified by sex and by O3I (≤5.6% vs >5.6%) (**Table 2**). Within sex comparisons found significant differences between individuals with O3I ≤5.6% vs. >5.6% for all variables in the adjusted model for females and males (**Table 2**). We observed a significant sex by O3I interaction in the adjusted model for RDW and BMI <u>for both sexes</u> with the difference in RDW below 5.6% being larger in females than males. <u>Across all O3I concentrations, RDW continued to decrease with increasing O3I in females whereas there was a plateauing in males, and even a modest increase in RDW as O3I exceeded 8%</u> (Figure 3).

**Table 2.** Characteristics of subjects by sex and omega-3 index (O3I) classification. (Mean±SD)

|  |  |  |  |  |  |
| --- | --- | --- | --- | --- | --- |
| Variable<sup>1</sup> | Female<sup>2</sup><br>(n=12,674) |  | Male<sup>3</sup><br>(n=12,811) |  | Sex*O3I Interaction |
|  | O3I ≤5.6%<br>(n=8,475) | O3I >5.6%<br>(n=4,199) | O3I ≤5.6%<br>(n=9,037) | O3I >5.6%<br>(n=3,774) | Adjusted<sup>4</sup><br>p-value |
| O3I (%) | 3.99±0.86 | 7.26±1.32 | 3.94±0.86 | 7.21±1.33 | 0.47 |
| RDW | 13.64±0.98 | 13.52±0.90 | 13.60±0.84 | 13.59±0.80 | 0.00066 |
| CRP (mg/L) | 1.31±0.79 | 1.13±0.75 | 1.26±0.76 | 1.09±0.71 | 0.03 |
| BMI (kg/m<sup>2</sup>) | 27.2±5.7 | 25.3±4.9 | 29.1±4.9 | 28.1±4.5 | <0.00001 |
| Age (years) | 52.6±15.6 | 57.9±13.5 | 52.6±14.6 | 57.9±13.3 | 0.90 |
| <sup>1</sup> O3I, omega-3 index; RDW, red blood cell distribution width; MCV, mean corpuscular volume; Hb, hemoglobin; CRP, high-sensitivity C-reactive protein; BMI, body mass index.<br><sup>2</sup> All variables between O3I categories (≤5.6% vs >5.6%) for females were p<0.0001 in both adjusted and unadjusted analyses.<br><sup>3</sup> All variables between O3I categories (≤5.6% vs >5.6%) for males were p<0.0001 in both adjusted and unadjusted analyses, except for unadjusted difference in RDW by O3I category, p=0.31.<br><sup>4</sup> Model further adjusted for age, BMI, and CRP. |  |  |  |  |  |

**Figure 2.**
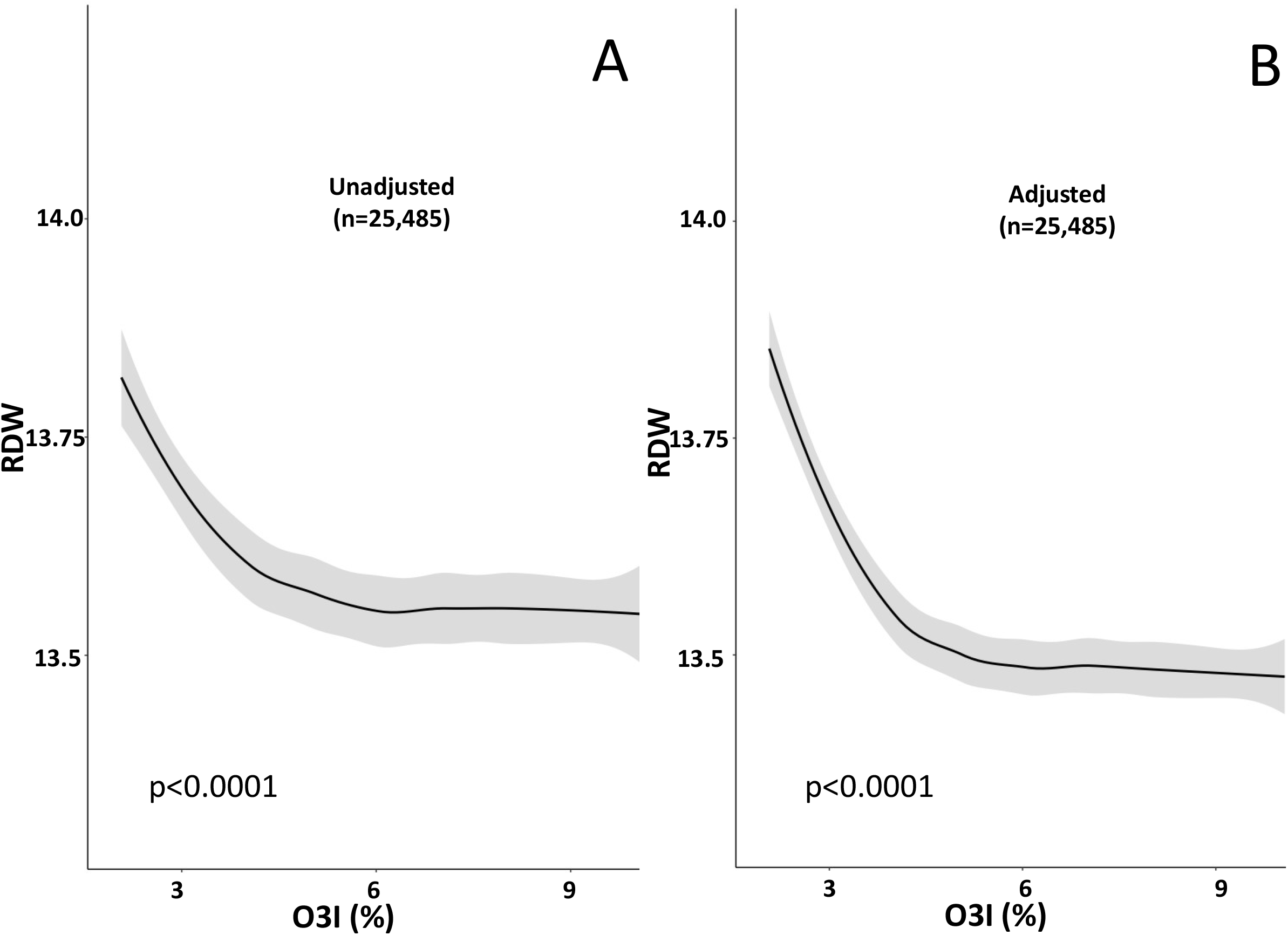
The unadjusted (A) and adjusted for age, sex, BMI and CRP (B) relationship between the red blood cell distribution width (RDW) and omega-3 index (O3I) in 25,485 adults without inflammation or anemia. (Predicted means and 95% confidence bands).

**Figure 3.**
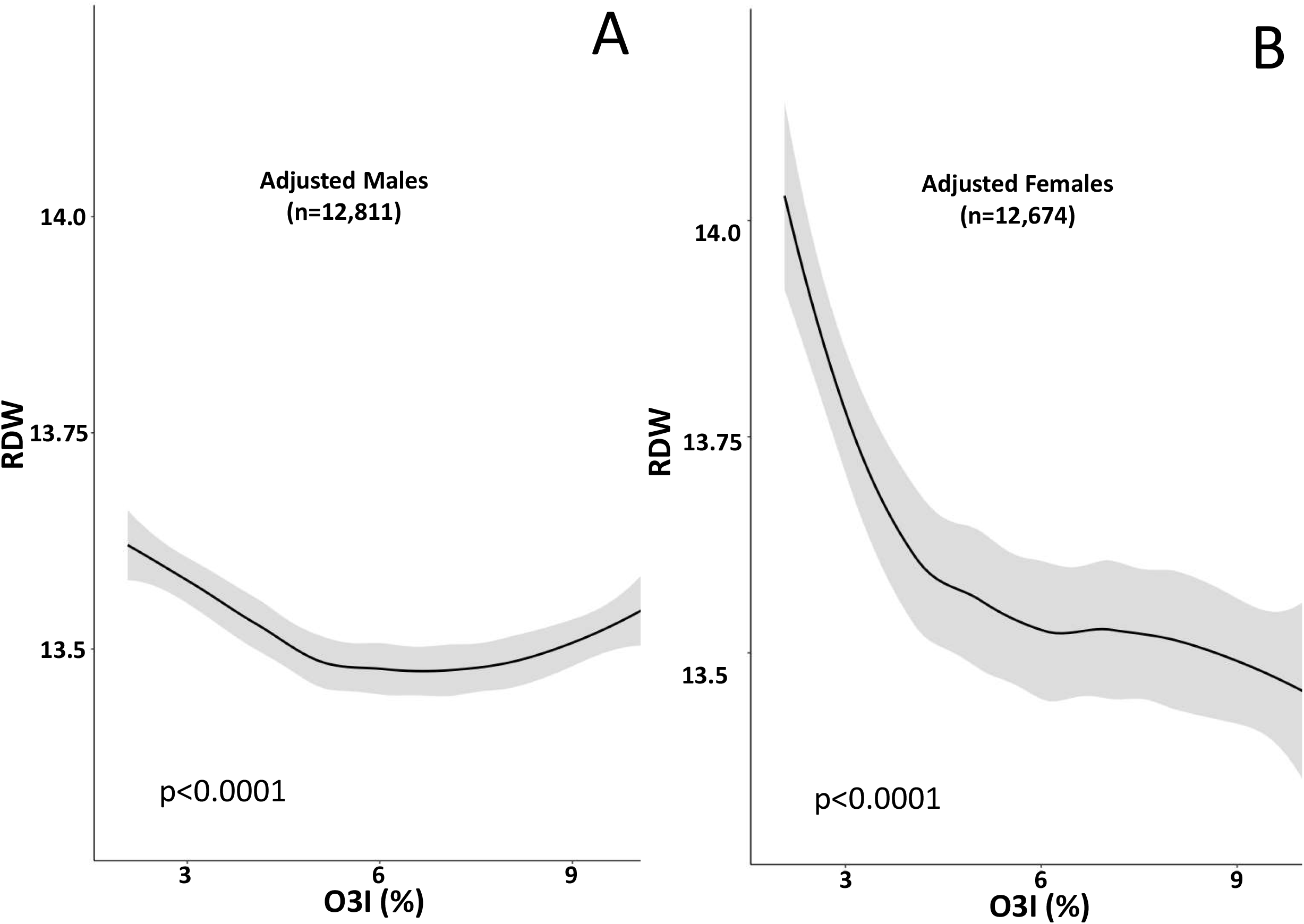
The unadjusted (A) and adjusted for age, BMI, and CRP (B) relationships between the red blood distribution width (RDW) and omega-3 index (O3I) in 12,811 males (A) and 12,674 females (B) without inflammation or anemia. (Predicted means and 95% confidence bands).

## 4. Discussion and Conclusions

RBCs are by far the most common cell in the body (83%), and as such, they have recently been viewed as a separate organ deeply involved with homeostasis [43]. Their primary role is to transport oxygen and carbon dioxide, and to accomplish this RBCs must pass through capillaries whose diameter is half of their own [44]. Thus, RBCs must be highly flexible and able to withstand high-shear stress. Here, we report an inverse relationship between the percentage of omega-3 fatty acids in RBC membranes and the distribution of RBC sizes in a generally healthy (i.e., not anemic and without inflammation) cohort (**Figure 2**), as well as an interaction with sex (**Figure 3**). <u>In both males and females, RDW was observed to be increasing with decreasing O3I below</u> 5.6% (**Figure 2; Table 2**). <u>The direction of the O3I-RDW relationship changed in males with O3I >∼8% but the increase in RDW is unlikely to have clinical significance.</u>

Changes in saturated and unsaturated fatty acid composition in cell membranes affect membrane fluidity [45–47] and deformability [28,48]. EPA and DHA have specific biophysical properties that affect the properties of membranes enriched in them [49]. Among these properties are fluidity, deformability, and susceptibility to aggregation of RBCs [26,28]. Increases in membrane DHA (a highly flexible molecule) influence the flexibility and structural integrity, i.e. lipid-protein interactions, of RBC membranes [50]. Increasing EPA intake alone or EPA+DHA together increases RBC membrane fluidity [48] and RBC deformability [25–27,51]. RBC deformability is affected by membrane lipid composition and lipid-protein interactions, and reduced deformability is a characteristic of aged RBCs [52]. Since an elevated RDW is likewise associated with decreased RBC membrane flexibility [2], one possible explanation for our findings is that a reduced EPA+DHA content of RBC membranes is causing the elevation in RDW.

An elevated RDW can also be caused by disordered erythropoiesis and/or RBC clearance [53]. A delay in clearance allows smaller RBCs to continue circulating, increasing the average age (lifespan) of RBC, and contributes to increased RDW [2]. Oxidative stress is a characteristic of aging, malnutrition, inflammation, and the pathogenesis of chronic disease [54]. Oxidative stress changes cytoskeletal arrangement and contributes to lipid asymmetry, causing erythrocytes lose flexibility and RDW to increase [55]. A systematic review and meta-analysis of 39 clinical trials (2,875 participants) found omega-3 supplementation improved antioxidant stability by significantly increasing serum total antioxidant capacity and decreasing serum glutathione peroxidase and malondialdehyde concentrations [56]. This represents another means by which omega-3 fatty acids could improve the RDW.

Alternatively, there may be a connection between omega-3, inflammation and the RDW. Inflammation marked by elevated CRP, interleukin-6, etc. can suppress RBC maturation in bone marrow, and larger immature RBCs, i.e. reticulocytes, enter the circulation and increase the RDW [16]. This relationship between RDW and inflammation may partially explain the relationship of higher RDW with risk of cardiovascular disease and mortality [10,11,14,30,32,57]. Similarly, lower omega-3 fatty acid levels are associated with a heightened inflammatory state [58–60], and omega-3 supplementation lowers inflammatory mediators [61]. Thus, a possible explanation of our findings may be that higher omega-3 fatty acid levels (O3I >5.6%) lower inflammatory markers which thereby lowers the RDW. <u>Because it has been recommended that a DRI be established for EPA+DHA</u> [62–65]<u>, the RDW-O3I analysis was purposefully constrained to healthy individuals with an objective of establishing a nutrient (EPA+DHA) structure-function (RDW) relationship. When the 14,239 individuals with inflammation (CRP >3 mg/L) were also included in the adjusted and unadjusted models, a similar inverse RDW-O3I relationship was determined (</u>**<u>Supplemental Figure 2</u>**).

Because of its cross-sectional nature, we cannot draw causal inferences from these data. However, there is evidence that raising RBC membrane omega-3 content can improve RDW. In patients with sickle cell disease, RBC rheological disturbances, hemolysis, and abnormalities of membrane fatty acids, have been ameliorated by omega-3 fatty acid treatment [66,67]. Supplementation with both EPA alone or EPA+DHA lowered RDW values in patients with coronary heart disease [51,68] and with abdominal aortic aneurysm [69]. In the latter study, supplementation increased O3I from ∼4.5% to 8% and decreased RDW from 14.8% to 13.8%. So, one of the mechanisms underlying the health benefits of EPA and DHA could be via an improved RDW.

The primary strength of this study was its large sample size which enabled us to explore RDW and O3I relationships in healthy individuals (males and females separately) free of laboratory signs of anemia or inflammation<u>, i.e.</u>, <u>a population suitable for the establishment of a DRI</u>. The most obvious limitations of this study, besides its cross-sectional nature, was the lack of information on confounding metabolic events, dietary behaviors, medications, other medical conditions and/or physical activities that might have also been associated with O3I and with RDW. Another potential limitation is that these data were drawn from a clinical laboratory database, not a formal national survey, so the representativeness of these patients to healthy US adults is not clear. However, previous studies in this specific cohort found that standard risk biomarkers (cholesterol, glucose, etc.) generally reflected those of typical adults living in the United States examined in Nutrition And Health Examination Surveys (NHANES) [70]. Also, O3I values were similar to those from a representative US sample [71], and RDW, MCV, and Hb were within normal ranges [7,72,73]. These considerations suggest this cohort was generally representative of the US population. The mean serum EPA+DHA concentration for adults 20+ years from NHANES was 2% [71] which is equivalent to an O3I of about 4.8%. Hence, roughly half of Americans have an O3I <5%, i.e., the range where higher RDW may be more common.

In conclusion, low O3I levels (≤5.6%) were associated with a potentially unhealthy distribution of RBC cell sizes. This suggests a role for EPA+DHA in maintaining RBC structure in healthy individuals. We propose that achieving an O3I >5.6% could help maintain a normal RBC size distribution. Our finding complements evidence that a higher O3I is linearly and inversely associated with risk for death from any cause [30], with values of >8% characterizing the lowest risk population [74]. Numerous researchers have called for the establishment of Dietary Reference Intake (DRI) for EPA+DHA [63–65]. To that end, our observations that maintaining an RBC EPA+DHA percentage above 5.6% may help maintain a normal RDW, could provide the scientific basis for a DRI.

## Data Availability

Pending application and approval, data described in the manuscript, code book, and analytic code will be made available upon request to the Fatty Acid Research Institute (https://www.faresinst.org/).

## Abbreviations

BMI: body mass index
CRP: high-sensitivity C-reactive protein
CV: coefficient of variation
DHA: docosahexaenoic acid
EPA: eicosapentaenoic acid
Hb: hemoglobin
MCV: mean corpuscular volume
O3I: omega-3 index
RBC: red blood cells
RDW: red blood cell distribution width
SD: standard deviation.

## Disclosures

M.I. McBurney serves on the Board of Directors of the American Society for Nutrition and has or has held consulting agreements in the past 3 years with Council for Responsible Nutrition; Church & Dwight; DSM Nutritional Products; International Life Sciences Institute, North America; McCormick; OmegaQuant Analytics; PepsiCo; and VitaMe Technologies. W.S. Harris holds an interest in OmegaQuant Analytics, a lab that offers omega-3 blood testing; and is a member of the RB Schiff Science and Innovation Advisory Board. N.L. Tintle has no conflicts to disclose.

## Funding

This work was supported by the Fatty Acid Research Institute (FARI). FARI is a non-profit foundation bringing together nutrition scientists and biostatistical experts to accelerate discovery of the relationships between fatty acids, especially omega-3 fatty acids, and health.

## CReditT authorship contribution statement

**M.I. McBurney**: Conceptualization & analytical design, Writing - original draft, Writing - review & editing. **W.S. Harris**: Conceptualization & analytical design, Funding - acquisition, Writing-review & editing. **N.L. Tintle**: Formal analysis, Writing - review & editing.

## Acknowledgements

The authors wish to thank Dr. Patrick Moriarty, Department of Internal Medicine, Kansas University Medical Center, Kansas City, KS for the initial suggestion to explore the relationship between omega-3 status and RDW. We also are indebted to Steven Varvel, PhD for his help in acquiring and collating the dataset used in this study.

## Notes

**Sources of Support:** This work was supported by the Fatty Acid Research Institute (FARI). FARI is a non-profit foundation bringing together nutrition scientists and biostatistical experts to accelerate discovery of the relationships between fatty acids, especially omega-3 fatty acids, and health. Pending application and approval, data described in the manuscript, code book, and analytic code will be made available upon request to the Fatty Acid Research Institute (https://www.faresinst.org/).

### Author Declarations

The Institutional Review Board of the University of South Dakota gave ethical approval for the analysis of deidentified laboratory data from HDL, Inc. (IRB-21-147).

### Summary of Updates

Low red blood cell (RBC) membrane content of EPA and DHA, i.e., the omega-3 index (O3I), and elevated RBC distribution width (RDW) are risk factors for all-cause mortality. O3I and RDW are related with membrane fluidity and deformability. Our objective was to determine if there is a relationship between O3I and RDW in healthy adults. Subjects without inflammation or anemia, and with values for O3I, RDW, high-sensitivity C-reactive protein (CRP), body mass index (BMI), age and sex were identified (n=25,485) from a clinical laboratory dataset of >45,000 individuals. RDW was inversely associated with O3I in both sexes before and after (both p<0.00001) adjusting models for sex, age, BMI and CRP. Stratification by sex revealed a sex-O3I interaction with the RDW-O3I slope (p<0.00066) being especially steep in females with O3I ≤5.6%. In healthy adults of both sexes, the data suggested that an O3I of >5.6% may help maintain normal RBC structural and functional integrity.

