## Supplemental Tables 1 and 2. Supp Figure 1 and 2. for "Omega-3 Index is Directly Associated with a Healthy Red Blood Cell Distribution Width"

[1]Fatty Acid Research Institute, Sioux Falls, SD 57106, USA (MIM, NLT, WSH); [2]Department of Human Health and Nutritional Sciences, University of Guelph, Guelph, ON N1G 2W1, Canada (MIM), [3]Division of Biochemical and Molecular Biology, Friedman School of Nutrition Science and Policy, Tufts University, Boston, MA 02111, USA (MIM); [4]Department of Population Health Nursing Science, College of Nursing, University of Illinois – Chicago, Chicago, IL 60612, USA(NLT); [5]Sanford School of Medicine, University of South Dakota, Sioux Falls, SD 57105, USA (WSH).

Supplemental Table 1. Red Blood Cell Distribution Width and the Omega-3 Index: Similar Relationships with Multiple Disease Outcomes.

| Condition or Outcome | Lower RDW* | | Higher Blood O3I | |
|---|---|---|---|---|
| | Setting | Finding | Setting | Finding |
| RBC Deformability | 293 healthy volunteers | Increased deformability (1) | Fish feeding trial | Improves (2) |
| RBC Aggregation | 105 SLE patients vs 105 controls | Reduced aggregation in SLE patients (3) | Fish feeding trial | Improves (2) |
| Inflammatory markers | 3845 outpatients | Lower biomarker levels (4) | Framingham | Lower biomarker levels (4) |
| CV Mortality | NHANES | Reduced risk (5) | FORCE meta-analysis | Reduced Risk (6) |
| Total mortality | | | FORCE meta-analysis | Reduced Risk (6) |
| Cancer mortality | Meta-analysis | Better outcomes (7) | FORCE meta-analysis | Reduced Risk (6) |
| Total mortality – patients with HF | Meta-analysis | Reduced risk (8) | No data | |
| Total mortality – patients with CHD | Meta-analysis | Reduced risk (8) | No data | |
| HF morbidity and mortality | 2679 CHF patients | Lower risk (9,10) | GISSI-HF trial<br>CHS observational study | Reduced mortality (11,12) and HF incidence (13) |
| Frailty | 619 Chinese mean age 69 y | Lower risk [15](14) | 1435 Koreans | Reduced risk (15) |
| Chronic kidney disease | 261 patients | Slower progression (16) | Observational study and O3I RCT in people with diabetes | Reduced risk (17) and slowed progression (18) |
| COVID-19 | 98 patients | Better outcomes (19,20) | 100 patients | Reduced risk for death (21) |
| Deep vein thrombosis/PE | Meta-analysis | Lower risk for PE (22,23) | Omega-3 RCT in post-operative fracture | Reduced risk for DVT (24) |

| | | | | |
|---|---|---|---|---|
| Atrial fibrillation | Swedish Health Records | Lower risk (25) | Danish National Cohort Adipose tissue omega-3 fatty acids | Reduced risk (26) |
| Metabolic syndrome | cross-sectional ~257,000 subjects | Lower odds (27) | Meta-analysis of omega-3 levels and incident T2DM | Lower risk (28) |
| Lupus | 105 SLE pts vs 105 controls | Less likely to be SLE patients (3) | RCT, 60 patients | Improved symptoms (29) |

*RDW, red blood cell distribution width; O3I, Omega-3 Index; RBC, red blood cell; SLE, systemic lupus erythematosus; CV, cardiovascular disease; FORCE, Fatty Acids and Outcomes Research Consortium; HF, heart failure; CHD, coronary heart disease; CHF; chronic heart failure; GISSI, Gruppo Italiano per lo Studio della Sopravvivenza nell'Infarto miocardioco Prevenzione trial; RCT, randomized controlled trial; DVT, deep vein thrombosis; T2DM, type 2 diabetes mellitus.

Supplemental Table 2. Characteristics of the total sample. Mean±SD

| Variable[1] | All (n=45,257) | Males (n=20,324) | Females (n=24,933) | Male vs Female Adjusted[2] p-value |
|---|---|---|---|---|
| O3I (%) | 4.80±1.76 | 4.77±1.77 | 4.82±1.75 | 0.005 |
| RDW | 13.86±1.22 | 13.80±1.10 | 13.91±1.30 | <0.00001 |
| MCV (fL) | 92.16±5.63 | 92.52±5.30 | 91.86±5.87 | <0.00001 |
| Hb[3] (g/dL) | 14.03±1.46 | 14.87±1.33 | 13.35±1.18 | <0.00001 |
| CRP[4] (mg/L) | 4.17±8.48 | 3.59±8.26 | 4.65±8.62 | <0.00001 |
| BMI (kg/m$^2$) | 29.4±6.7 | 29.8±5.8 | 29.1±7.4 | <0.00001 |
| Age (years) | 55.1±15.1 | 55.6±14.8 | 54.6±15.4 | <0.00001 |

[1]O3I, Omega-3 index; RDW, red blood cell distribution width; MCV, mean corpuscular volume; Hb, hemoglobin; CRP, high-sensitivity C-reactive protein; BMI, body mass index.

[2]Models were adjusted for age, BMI, and CRP (except when the model was predicting these variables).

[3] 258 individuals were missing Hb values.

[4] 1098 individuals were missing CRP values.

Online Supplemental Figure 1. The adjusted for age, sex, BMI and CRP relationship between the red blood cell distribution width (RDW) and eicosapentaenoic acid (A), docosahexaenoic acid (B), and arachidonic acid (C) content (%) in 25,485 healthy adults without anemia or chronic inflammation. (Predicted means and 95% confidence bands).

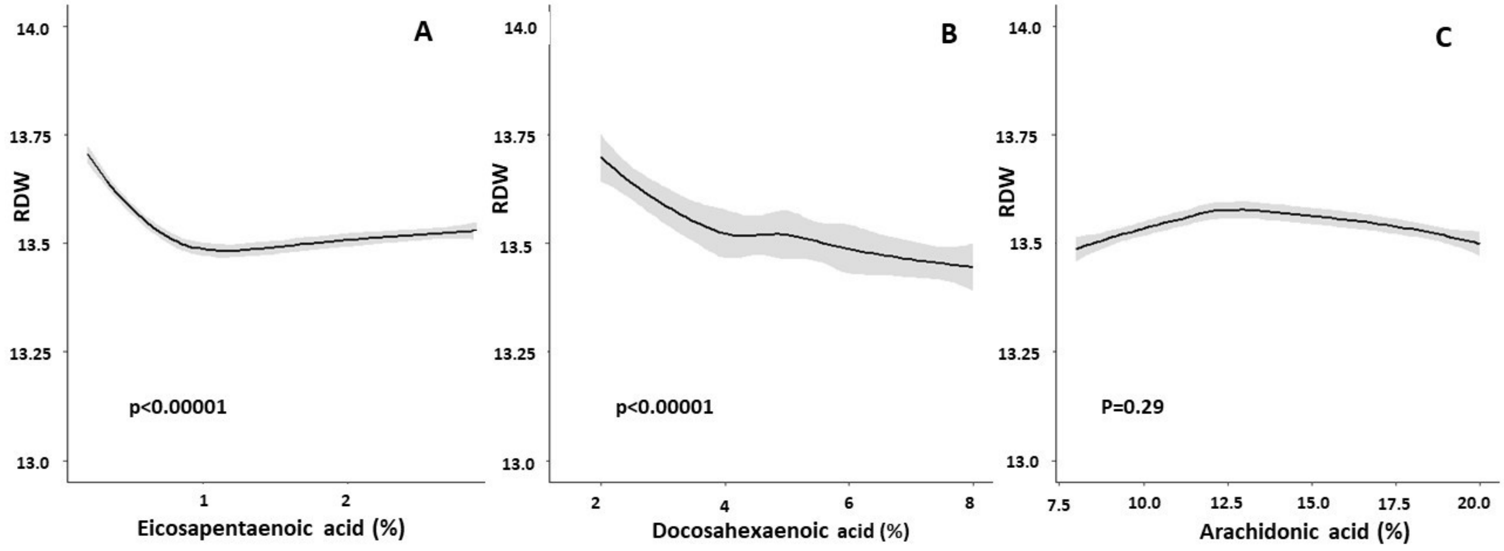

Online Supplemental Figure 2. The unadjusted (A) and adjusted for age, sex, BMI and CRP (B) relationship between the red blood cell distribution width (RDW) and omega-3 index (O3I) in 45,257 adults (includes 2,480 individuals with anemia, 1,055 individuals missing CRP values, and 3,818 individuals with acute inflammation (CRP> 10 mg/L)). (Predicted means and 95% confidence bands).

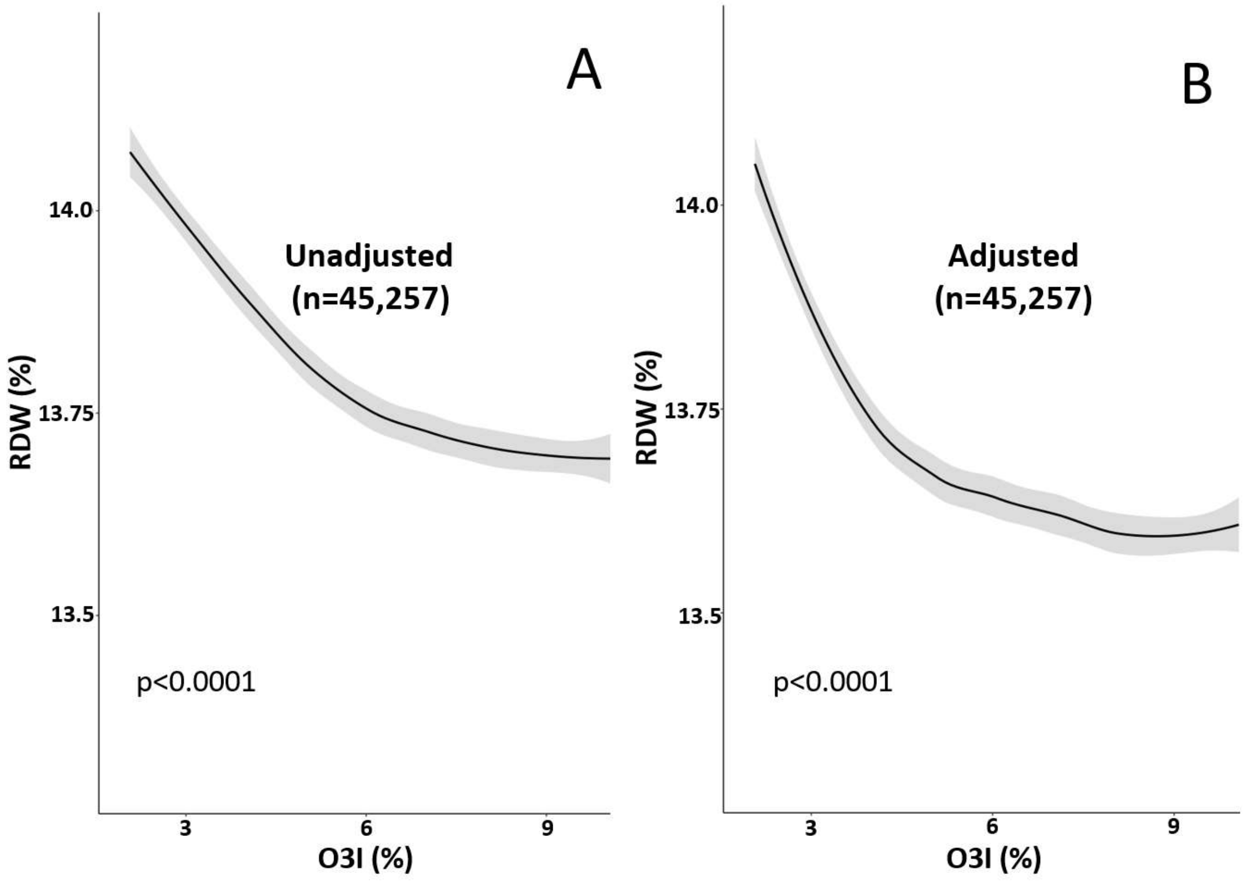

10. Allen LA, Felker GM, Mehra MR, Chiong JR, Dunlap SH, Ghali JK, Lenihan DJ, Oren RM, Wagoner LE, Schwartz TA, et al. Validation and Potential Mechanisms of Red Cell

Distribution Width as a Prognostic Marker in Heart Failure. Journal of Cardiac Failure 2010;16:230–8.

20. Rapp JL, Tremblay D, Alpert N, Lieberman-Cribbin W, Mascarenhas J, Taioli E, Ghaffari S. Red cell distribution width is associated with mortality in non-anemic patients with COVID-19. J Med Virol 2021;jmv.27011.
